# A factorial Mendelian randomization study to systematically prioritize genetic targets for the treatment of cardiovascular disease

**DOI:** 10.1101/2020.02.16.20023010

**Authors:** Genevieve M. Leyden, Tom R. Gaunt, Tom G. Richardson

**Affiliations:** MRC Integrative Epidemiology Unit, University of Bristol; Bristol Medical School: Translational Health Sciences, University of Bristol

**Author notes:** Corresponding author: Dr. Tom G. Richardson, MRC Integrative Epidemiology Unit, Bristol Medical School (Population Health Sciences), University of Bristol, Oakfield House, Oakfield Grove, Bristol BS8 2BN, United Kingdom.

**Keywords:** Mendelian randomization, factorial study design, drug validation, phenome-wide association study, UK Biobank

## Abstract

**Importance:** New drugs which provide benefit alongside statin mono-therapy are warranted to reduce risk of cardiovascular disease.

**Objective:** To systematically evaluate the genetically predicted effects of 8,851 genes and cardiovascular disease risk factors using data from the UK Biobank and subsequently prioritize their potential to reduce cardiovascular disease in addition to statin therapy.

**Design, Setting, and Participants:** A factorial Mendelian randomization study using individual level data from the UK Biobank study. This population-based cohort includes a total of 502,602 individuals aged between 40 and 96 years old at baseline who were recruited between 2006 to 2010.

**Exposures:** Genetic variants robustly associated with the expression of target genes in whole blood (based on P<5×10^−08^) were used to construct gene scores using findings from the eQTLGen consortium (n=31,684).

**Main Outcomes and Measures:** Genetically predicted effects for each of the 8,851 genes were firstly evaluated with 5 measured outcomes from the UK Biobank in turn (body mass index, diastolic blood pressure, systolic blood pressure, low-density lipoproteins and triglycerides). Effects surviving multiple comparisons from this initial analysis were subsequently analyzed using factorial Mendelian randomization to evaluate evidence of an additive beneficial effect on cardiovascular disease risk compared to a *HMGCR* genetic score acting as a proxy for statin inhibition. Finally, a phenome-wide analysis was undertaken to evaluate predicted effects between prioritized targets and 569 outcomes in the UK Biobank to highlight potential adverse side-effects.

**Results:** 377 genetically predicted effects on cardiovascular disease risk factors were identified by Mendelian randomization (based on P<1.13×10^−6^). Of the 68 druggable genes, 20 candidate genes were predicted to lower cardiovascular disease risk in combination with statin treatment (P<7.35×10^−4^). Genes such as *FDFT1* were predicted to have added benefit with statin therapy (OR=0.93; 95% CI, 0.91-0.95; P=2.21×10^−10^) but are unlikely to make safe targets due to their predicted effects on adverse side effects. In contrast, *PRKCE* provided evidence of a putative added benefit in combination with statins (OR=0.94; CI, 0.91-0.96; P=1.72×10^−9^) with no predicted adverse effects.

**Conclusions and Relevance:** Through the application of a systematic factorial Mendelian randomization analysis, we have prioritized (and deprioritized) potential drug targets predicted to reduce cardiovascular disease risk in addition to statin therapy.

**Key points:** *Question:* Can naturally occurring genetic variation in a population help us highlight and prioritize novel therapeutic targets for the treatment of cardiovascular disease?

*Findings:* In this factorial Mendelian randomization study of 334,915 individuals, we found that a genetically predicted 0.09 mmol/L decrease in LDL cholesterol attributed to statin inhibition results in 4.1% lower risk of cardiovascular disease. We then highlighted various genetic targets which were genetically predicted to further reduce disease risk without evidence of adverse side effects, such as *PRKCE* which is involved in the development of cardiac hypertrophy and reduced risk of cardiovascular disease by 6.4% in addition to statin therapy.

*Meaning:* Evidence from genetic analyses can improve the likelihood of success for therapeutic targets and findings from this study have prioritized several promising candidates for the treatment of cardiovascular disease.

## Introduction

Cardiovascular disease (CVD) is an increasingly prevalent public health concern and remains the leading cause of death worldwide ^1^. Cholesterol lowering drugs such as statins (3-hydroxy-3-methylglutaryl coenzyme A reductase [*HMGCR*]-inhibitors) are regarded as the gold-standard treatment option in terms of lowering the risk of CVD including myocardial infarction and stroke^2,3^. Whilst the effectiveness of statins in risk reduction for both primary and secondary CVD has been established in randomized control trials (RCTs) ^4,5^, they have also been reported to have adverse side-effects in certain patients, such as an increased risk of developing type-2 diabetes ^6^ and weight-gain ^7^. Furthermore, there is major unmet clinical need for the identification of additional drugs to achieve adequately lower CVD risk in patients undergoing statin mono-therapy ^8^ or a viable alternative to it ^9^.

Mendelian Randomization (MR) is a technique in causal inference which uses naturally occurring genetic variation to investigate the relationship between modifiable exposures (such as the anticipated effect of a drug) and disease outcomes ^10,11^. By exploiting the random assortment of genetic alleles at birth, MR is often considered analogous to the allocation of individuals to drug and placebo groups in an RCT, without the concerns of non-adherence^12^. As such, findings from MR are less prone to confounding and reverse causation which can hinder classical observational studies.

Recent studies have demonstrated the value of conducting genetic and MR analyses to mimic the putative effects of therapeutic intervention ^13-15^. Multiple studies have examined the relative relationship between lifelong genetic inhibition of *HMGCR* and alternative genetic targets ^16,17^. These studies have used a 2×2 factorial approach, which stratifies the sample population by allelic risk scores to estimate the separate and combined effects of genetic proxies of therapeutic intervention on CVD outcomes. Such developments have established MR as a powerful approach for drug discovery and improved understanding of disease aetiology.

In this study, we describe an MR framework designed to systematically prioritize putative genetic targets which are predicted to have an independent and additive benefit alongside statin treatment. This approach was applied to evaluate genetically predicted effects of 8,851 genetic targets on measures of CVD risk in the UK Biobank study (UKB) ^18^. Genes identified in this analysis were subsequently analyzed using 2×2 factorial to assess whether therapeutically targeting them may have a predicted reduced risk on CVD in addition to statin therapy. Finally, we performed a phenome-wide association study (PheWAS) to highlight any putative adverse effects for prioritized targets identified in the previous analyses. In doing so, we demonstrate the ability of this framework to capture diverse biological functions and recapitulate results outlined in preclinical studies, strengthening the validity of this approach to prioritize (and deprioritize) drug targets for therapeutic validation.

## Methods

### Study populations and outcomes

Single nucleotide polymorphisms (SNPs) robustly associated with changes to gene expression (i.e. P<5×10^−08^) were selected as instrumental variables using findings from the eQTLGen consortium (n=31,684) ^19^. Our inclusion criterion was based on genes whose expression could be instrumented by at least 2 independent SNPs within a 1Mb distance of a gene’s transcription start site (known as cis-eQTL). This was to improve the robustness of findings in line with the assumptions of MR, further details of which can be found in supplementary methods. A reference panel of European individuals from the 1000 genome project (phase 3) was used to identify independent SNPs based on r^2^<0.01 ^20^.

The UK Biobank is a prospective cohort study with detailed genotype and phenotype data on up to 500,000 participants ^18^. The CVD risk factors we evaluated in our initial analysis using data from UKB were; body-mass index (BMI), diastolic blood pressure (DBP), systolic blood pressure (SBP), low-density lipoproteins (LDL) and triglycerides (TG). A derived outcome encompassing all CVD outcomes from UKB using data from field 20002 (such as coronary heart disease, hypertension and hypercholesterolemia) was used in the factorial 2×2 MR analysis. Individuals were therefore categorized as a case in our derived variable if they were a case for any CVD outcomes in this field. The phenome-wide analysis used data on 569 outcomes from UKB as described previously ^21^, a full list of which can be found in Supplementary Table 11.

### Statistical analysis

#### Identifying associations between genetically predicted gene expression and risk factors for cardiovascular disease

To assess whether changes in gene expression have a putative causal role in CVD risk, cis-eQTL were harnessed as instrumental variables in a 2-sample MR analysis ^22^. We applied the inverse variance weighted method (IVW) ^23^ to estimate the effect of genetically predicted gene expression on each of the 5 CVD traits in turn using the ‘TwoSampleMR’ package ^24^. Genetically predicted effects which survived multiple testing (based on P < 0.05/(number of genes analyzed*number of traits)) were subsequently filtered to identify those which were candidates for therapeutic intervention and therefore carried forward to downstream analyses. Targets were also filtered to identify those which were “druggable” using a comprehensive list of genes collated from recent data-driven drug-discovery and target selection strategies ^25-28^.

#### Systematic factorial Mendelian randomization analysis to prioritize therapeutic targets

Individual-level genetic data was used from participants in UKB to construct genetic risk scores (GRS) for each ‘druggable’ gene associated with any of the 5 traits from the previous analysis. GRS were constructed as the sum of the effect alleles for eQTL SNPs weighted by their eQTLGen regression coefficients. To account for multiple testing in the factorial analysis, the genetically predicted effect between each GRS and CVD in UKB (based on data from field 20002 as described above) was estimated by using logistic regression with adjustment for age, sex and the first 10 principal components (PC) of ancestry. Only genes whose predicted effect on the derived CVD outcome (based on a multiple testing correction of P<0.05/number of GRS evaluated) were analyzed in the factorial MR analysis. A previously published *HMGCR* score was constructed using 6 LDL-associated variants within 100kb of this gene to mimic the CVD lowering effect on statin therapy ^16^.

We applied 2×2 factorial MR to systematically compare the effect of each gene associated with CVD in the previous analysis after accounting for the effect of the *HMGCR* score. The UKB dataset consisting of 334,915 individuals (described in further detail in the supplementary methods) was first dichotomized into halves based on the median of the *HMGCR* score, and then into quarters based on the median of the potential novel gene target’s GRS. The group consisting of individuals with below median values for both the *HMGCR* and novel gene scores was subsequently used as the baseline group in further analyses. Each of the other groups were analyzed in turn with the baseline group to estimate the effect of genetically predicted effects for statin inhibition, new drug effect and a combined therapy. A graphical illustration of this approach can be found in Figure 1. Analyses involved logistic regression on the CVD outcome adjusting for the same covariates as before.

**Figure 1.**
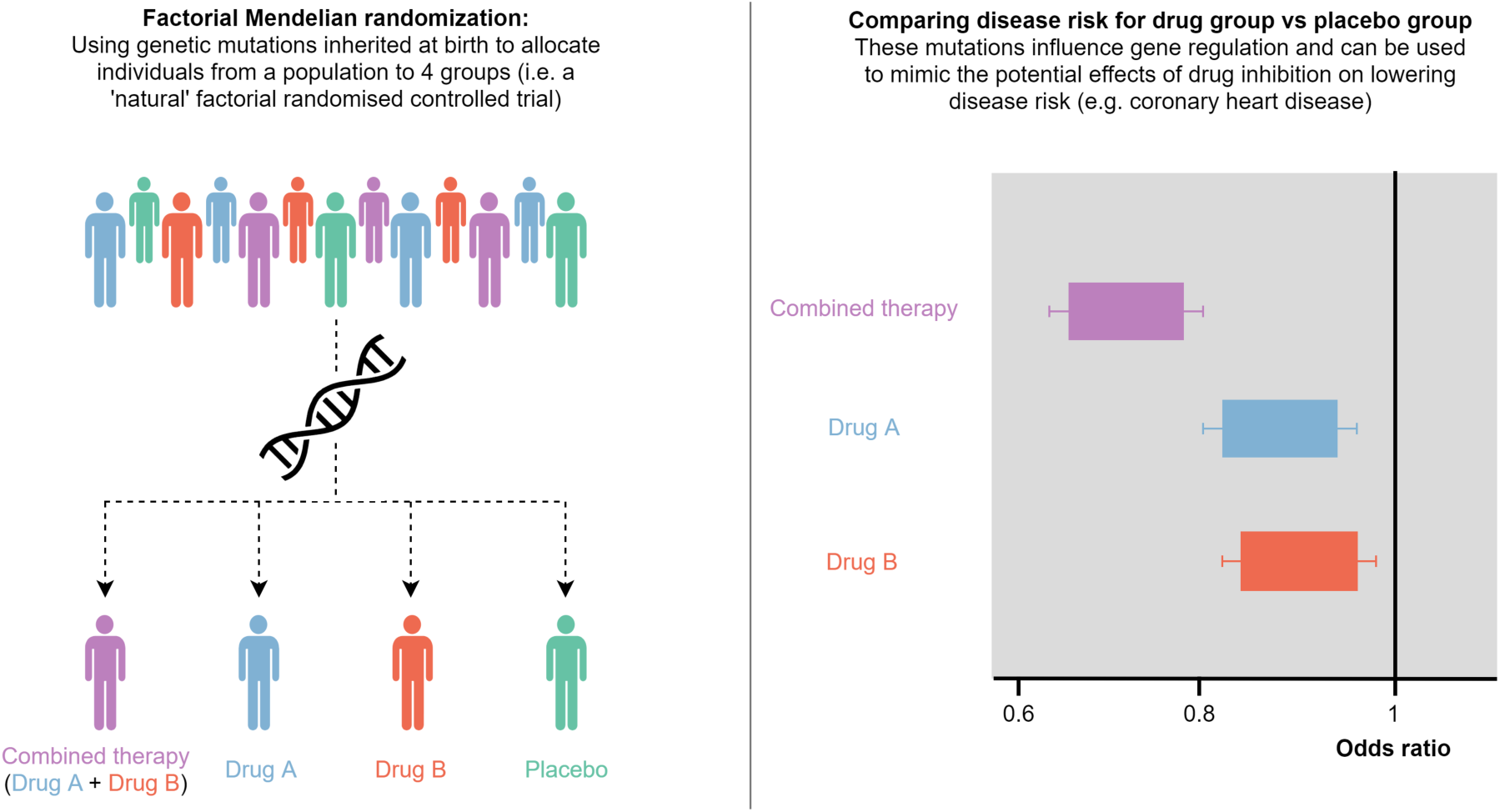
An illustration of the concept behind factorial Mendelian randomization. Using naturally occurring genetic mutations in a population, individuals are allocated to 4 different groups depending on whether they harbor mutations known to influence the regulation of two target genes. These mutations are combined into two scores, one for each gene, which can be used to mimic the potential impact of inhibiting their regulation. As such, people allocated to the placebo group have a score less than the median for both genes, Drug A/Drug B groups have a score higher than the median for one gene but not the other, whereas those in the combined therapy group have higher scores than the median for both genes. By comparing incidence of disease in each group with the placebo group, it is possible to infer whether developing a drug for a novel gene target would yield an additive therapeutic benefit over current treatments. We have demonstrated this in our study to assess whether evaluating novel drug targets may be worthwhile in terms of treating coronary heart disease on top of *HMGCR* inhibition (i.e. statin therapy).

#### Phenome-wide association study (PheWAS)

Finally, we undertook a PheWAS analysis for each GRS prioritized in the previous analyses to highlight any potentially unanticipated adverse effects of therapeutic intervention. This was undertaken by evaluating the association between each GRS and 569 outcomes from the UKB. Continuous, binary and categorical traits were analyzed using linear, logistic and ordinal/multinomial logistic regression respectively. All analyses were adjusted for age, sex and the top 10 PCs. All analyses were undertaken using R (version 3.5.1) and all plots were created using the package ‘ggplot2’.

## Results

### A systematic Mendelian Randomization analysis to identify novel candidate genes associated with cardiovascular disease

The putative causal effect between the genetically predicted expression of 8,851 genes (Supplementary Table 1) and 5 cardiovascular disease traits in UKB (BMI, SBP, DBP, LDL and TG) was systematically screened in 44,255 distinct MR analyses. Overall, 377 genetically predicted effects were identified which survived the Bonferroni corrected threshold (Bonferroni P<0.05/44,255=1.13×10^−6^) (Figure 2, Supplementary Table 2-6). A flow diagram summarizing downstream findings in this study can be found in Figure 3.

**Figure 2.**
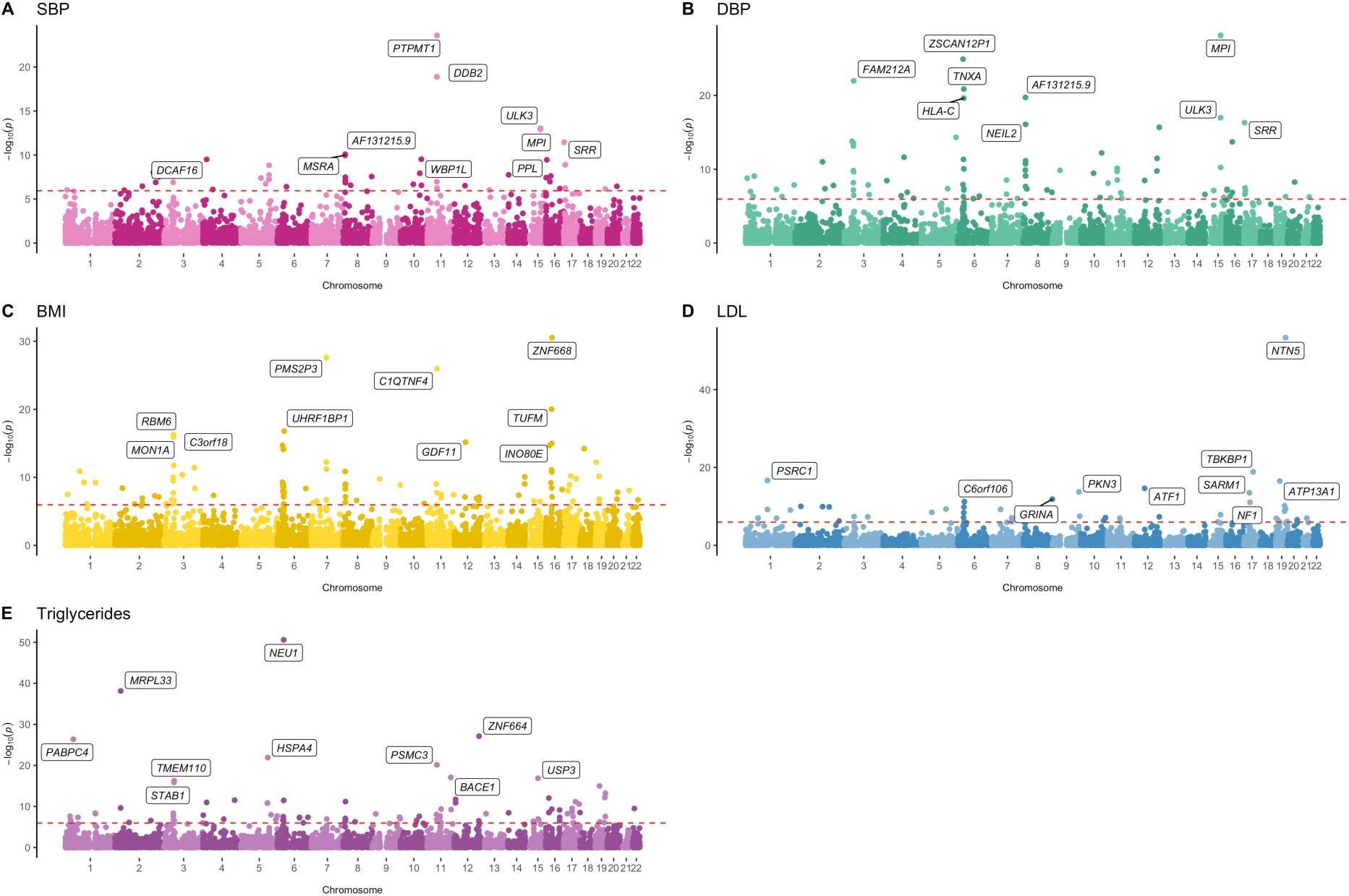
Manhattan plots illustrating the associations between the expression of 8,851 genes and 5 known risk factors for cardiovascular disease. The y-axis indicates the -log^10^ p-value for each genetically predicted effect with the corresponding gene’s genomic location on the x-axis. Gene annotations refer to the top 10 predicted effects per trait. The horizontal dashed red line indicates the Bonferroni multiple testing threshold in this analysis (i.e. P<0.05/(8,851*5) = 1.1298×10^−6^). Results for systolic blood pressure, diastolic blood pressure, body-mass index, LDL-cholesterol, and triglycerides are plotted on plots A-E respectively.

**Figure 3.**
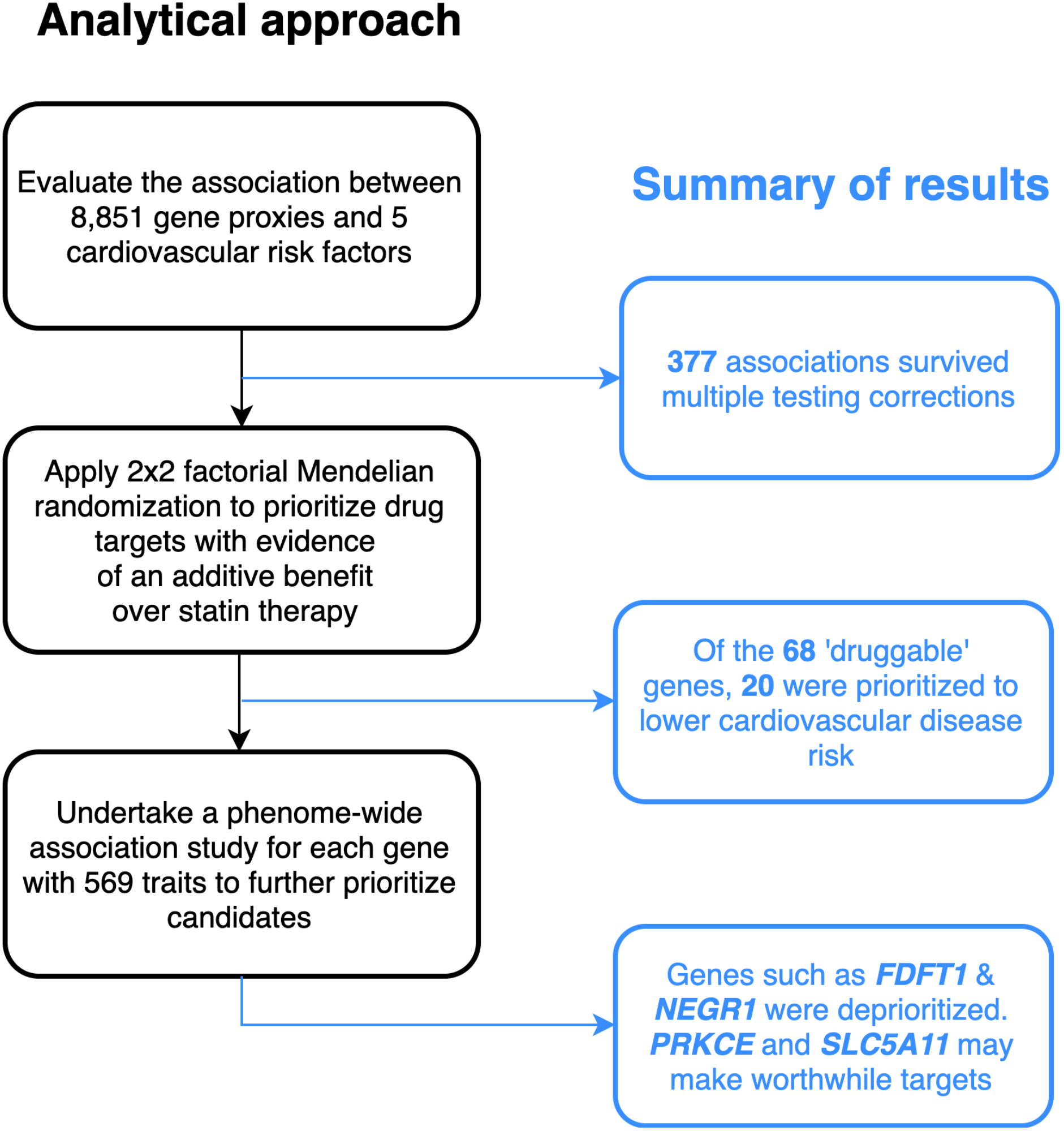
Flow diagram of study analysis. A flow diagram illustrating the different stages of this study and a summary of findings.

Leveraging multiple SNPs in an MR analysis may provide additional power to detect associations that may previously have been overlooked by conventional single SNP analyses. Filtering for genes whose individual SNP instruments would not have survived genome-wide corrections (i.e. P>5×10^−8^), yet their combined IVW estimates resulted in an effect which would have (P<5×10^−8^), indicated that 67 of these association signals (17.8%) would potentially have been missed by conventional single SNP analyses (Supplementary Table 7). For example, the association between *FADS1* and triglycerides would have been identified in a GWAS based on the strongest individual SNP effect for this gene (P=5.5×10^−109^). However, the association between *POM121C* would have been overlooked by conventional single SNP analyses (lowest individual P=9.40×10^−5^), yet the combined effect of all eQTL provided stronger evidence of association (IVW P=5.83×10^−13^). *POM121C* encodes a nucleoporin whose expression may be involved in regulating insulin sensitivity and adipogenesis ^29^.

There were 68 candidate genes of the 377 putative effects that were “druggable”, as defined by recent data-driven drug-discovery and target selection strategies ^25-28^ (Supplementary Table 8). Of these, there was strong evidence of a genetically predicted effect between the expression of 20 genes and the CVD outcome in UKB based on our GRS analysis (Bonferroni P=0.05/68=7.35×10^−4^).

### Prioritizing cardiovascular genes for therapeutic intervention using factorial Mendelian randomization

Next, we sought to discern whether there was genetic evidence of an additive CVD risk lowering effect of the 20 identified candidate genes compared to an *HMGCR* score acting as a proxy for statin inhibition (Supplementary Table 9). The *HMGCR* score was strongly associated with LDL cholesterol (Beta=-0.09, SE=0.004, P=5.88×10^−115^) and CVD (OR=0.96, 95% CI=0.95-0.97, P=8.81×10^−05^) in the UK Biobank study. This suggests that each 0.09 mmol/L decrease in LDL cholesterol attributed to statin inhibition results in 4.1% lower risk of CVD based on these estimates.

A summary of existing compounds for these genetic targets and their indications is provided in Supplementary Table 10. The genes with the largest magnitude of an additive effect included *FDFT1*, which encodes squalene synthase (SQS), a protein downstream *HMGCR* on the cholesterol biosynthesis pathway ^30^. Individuals in the *FDFT1-HMGCR* “combined” low-risk group were predicted to have lower odds of developing CVD (OR=0.93 per standard deviation increase in GRS; 95% CI, 0.91-0.95; P=2.21×10^−10^) than the “statin” group (OR=0.97; CI, 0.95-0.99; P=0.0053) (Figure 4). Similarly lower CVD risk estimates were predicted for the *NEGR1* combined effect with statins (OR=0.93; CI, 0.91-0.95; P=2.80×10^−11^) than the statin only group (OR=0.97; CI, 0.95-0.99; P=0.0068), and for *PRKCE*-combined (OR=0.94; CI, 0.91-0.96; P=1.72×10^−9^) vs statin only (OR=0.965; CI, 0.94-0.99; P=0.0014). In comparison, genes such as *SLC5A11* were associated with a more modest decreased risk of CVD in the “combined” (OR=0.94; CI, 0.92-0.97; P=1.13×10^−7^) than the “statin”-group (OR=0.96; CI, 0.94-0.98; P=8.52×10^−5^) (Figure 4).These analyses therefore suggest that the benefit of combined therapy (i.e. statins + new drug) for these novel gene targets would result in a reduction of between 6-7% reduction in CVD risk over the placebo group.

**Figure 4.**
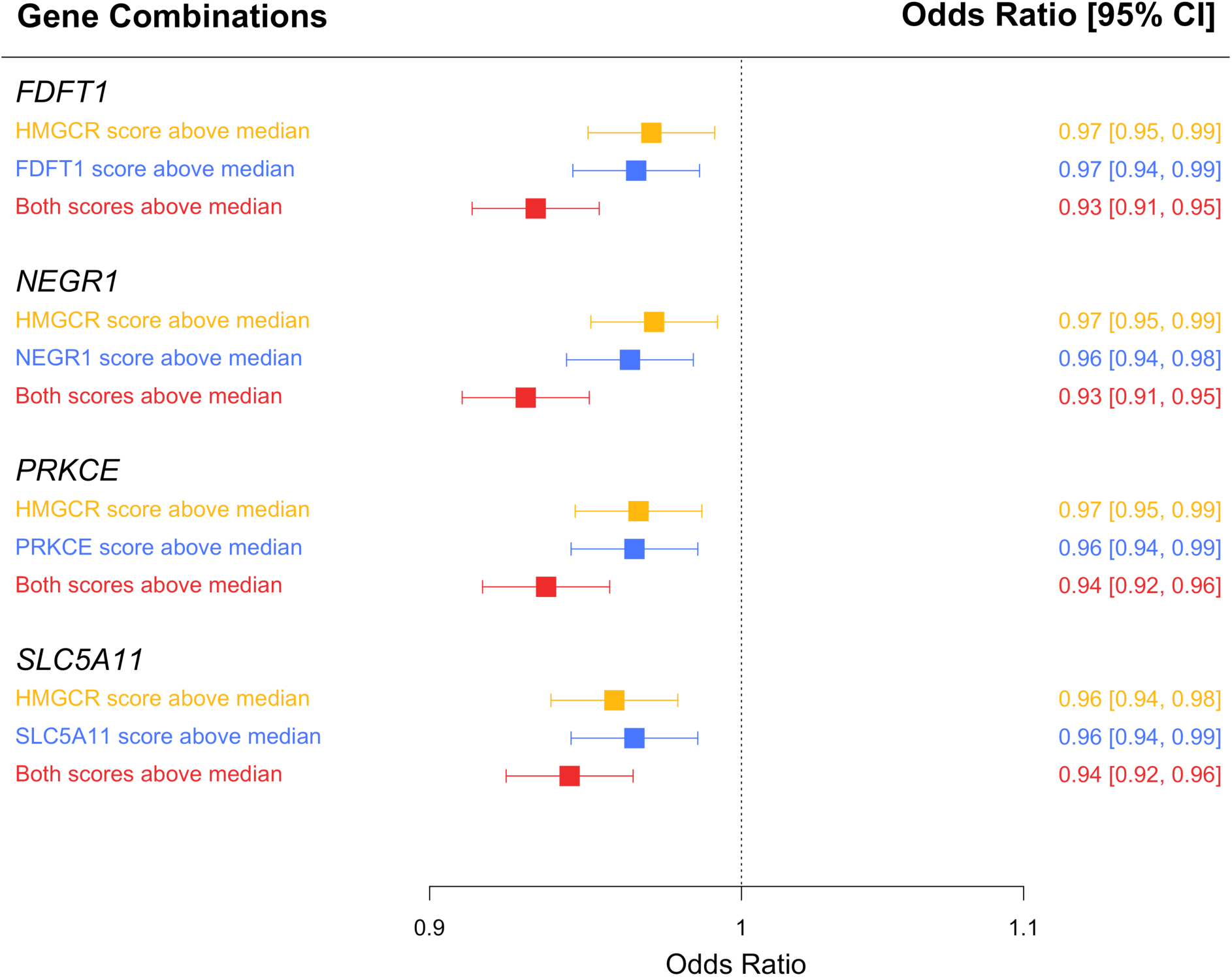
A forest plot to illustrate 2×2 Factorial Mendelian Randomization estimates. A comparison of findings from the 2×2 factorial Mendelian randomization analysis for 4 drug targets (*FDFT1, NEGR1, PRKCE* and *SLC5A11*). Genetic risk scores for each of these genes were constructed in the UK Biobank cohort and evaluated against a previously devised score for *HMGCR* by Ference et al (2015). In turn, each score was compared with the *HMGCR* score to assess whether targeting them may lower cardiovascular disease risk in addition to statin therapy.

### Exploring phenome-wide associations to predict putative side effects of genetically targeted therapeutics

As a proof of concept, we firstly undertook a PheWAS to evaluate the genetically predicted effects of *HMGCR* using the score used in the previous analysis (Figure 5A, Supplementary Table 12). The results predict that *HMGCR* is associated with increased cholesterol (β= 0.038 per standard deviation increase in GRS; 95% CI, 0.035-0.041; P=1.36×10^−106^) and LDL cholesterol (β= 0.040; CI, 0.036-0.043; P=7.92×10^−113^). As such, inhibiting *HMGCR* will have a predictively beneficial lowering effect on these traits which is the anticipated outcome of statins. For example, statin use has been reported to increase risk of type-2 diabetes which is likely reflected in the PheWAS results which identified an association with increased glycated haemoglobin (HbA1c) (β= −0.008; CI, −0.012--0.004; P=4.10×10^−6^), a diagnostic marker of type-2 diabetes.

**Figure 5.**
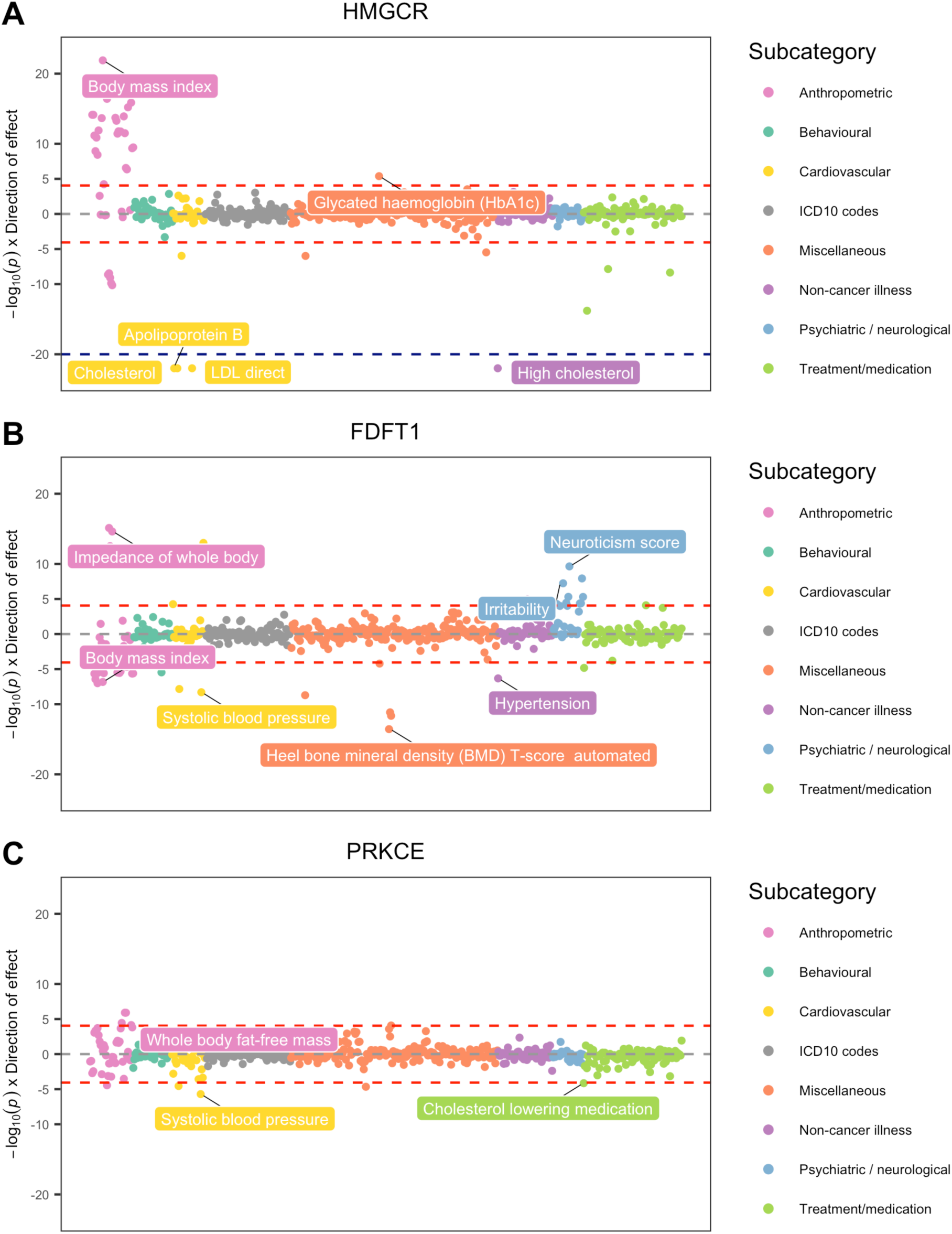
Phenome-wide association plot for predicted genetic inhibition of (A) *HMGCR*, (B) *FDFT1* and (C) *PRKCE*. Each point on the plot represents the association between the respective genetic score and a complex trait in UKB. The y-axis indicates the -log10 p-value for each association after orienting their direction of effect in line with predictive therapeutic inhibition (i.e. statins have an LDL cholesterol lowering effect and as such they reside below the null). Points are grouped and colored according to the corresponding subcategory for each trait. The horizontal dashed red line indicates the Bonferroni corrected threshold for 569 traits (i.e. P=-log10(8.79×10^−5^).

Full results from the PheWAS analyses for each of the 20 genes taken forward from the previous analysis can be found in Supplementary Tables 13-32). As expected, the PheWAS results for prioritized genes indicated an enrichment for traits with known roles in mediating CVD risk, in addition to novel secondary effects which may raise concern for the development of compounds targeting multiple pathways. For instance, *FDFT1* and *NEGR1* were associated with 55 and 50 traits respectively (Bonferroni P=0.05/569=8.79×10^−5^) (Figure 5B, Supplementary Figure 1A; Supplementary Table 19 & 27). In addition to CVD related outcomes, there were also genetically predicted effects which may foreshadow adverse effects. For example, *FDFT1* expression was associated with alkaline phosphatase (β= 0.022; CI, 0.018 - 0.022; P=1.41×10^−35^) and alanine aminotransferase (β= 0.010; CI, 0.006 - 0.013; P=1.05×10^−08^) which are markers of liver disease^31^. There was also evidence that *NEGR1* expression associates with fluid intelligence score (β= 0.026; CI, 0.014-0.039; P=3.69×10^−5^) in the same direction to anthropometric traits, suggesting that inhibiting *NEGR1* may have deleterious consequences on this trait.

In contrast, findings from the PheWAS analysis suggested that alternative targets may provide more viable therapeutic opportunities for genes such as *SLC5A11* and *PRKCE* (Figure 5C, Supplementary figure 1B, Supplementary Tables 28 & 30). *SLC5A11* was associated with anthropometric traits such as BMI (β= −0.01; CI, −0.013--0.007; P=3.82×10^−9^), as well as cardiovascular traits including SBP (β= −0.012; CI, −0.015--0.008; P=1.63×10^−12^). *PRKCE* was more specifically associated with cardiovascular traits influencing blood pressure traits such as SBP (β=0.008; CI, 0.005-0.011; P=1.99×10^−6^) and DBP (β= 0.007; CI, 0.004-0.01; P=2.83×10^−5^). Notably, there were no associations which provided strong evidence of potentially adverse secondary effects in this analysis for either gene.

## Discussion

In this study we present a comprehensive analytical pipeline which harnesses Mendelian randomization to assess whether the inhibition of novel drug targets may further reduce risk of cardiovascular disease in addition to statin treatment. Applying this framework in the UK Biobank study prioritized 20 genetic targets which were predicted to provide additional therapeutic benefit in combination with statins, including known cooperative drug interactions ^32^. Exploring the putative effects of these targets on 569 outcomes supported the validity of this methodology at estimating potential secondary drug effects. For example, the *HMGCR* score was strongly associated with HbA1C in the opposite direction to LDL cholesterol, which is indicative of the increased risk of type 2 diabetes which may accompany statin therapy based on clinical evidence^33^. This approach was subsequently applied to each of the 20 identified genes detected to further prioritize their potential as a therapeutic target.

The use of large-scale genotype-phenotype datasets is becoming increasingly important as an early drug-development tool for informed target validation ^34^. Our two-sample MR analysis using genetic instruments which influenced the expression of genetic targets identified 377 putative effects across 5 CVD risk factors. Amongst these findings were several candidate genes that have been evaluated previously in pre-clinical and clinical studies. This includes *FDFT1* which encodes squalene synthase (SQS), a protein downstream *HMGCR* on the cholesterol biosynthesis pathway ^30^. The therapeutic potential of SQS-inhibition has been explored in preclinical studies demonstrating cooperation between SQS-inhibitors and statins in achieving increased LDL-cholesterol clearance ^35^. Despite our findings supporting the efficacy of *FDFT1*, there was also evidence of putative adverse side-effects. This included genetically predicted effects on alkaline phosphatase and alanine aminotransferase which suggests that this targets’ protein product may have downstream consequences for liver function. SQS-inhibitors have been developed to late-stage clinical trial but have been discontinued due to treatment associated hepatotoxicity ^36^.

Similarly, while the factorial MR analyses provided evidence that *NEGR1* may be an effective therapeutic target in lowering CVD risk in combination with statin treatment, its predicted effects on neurological/psychiatric traits may complicate its specificity. *NEGR1* (neuronal growth regulator-1) is expressed in the hypothalamus plays a role in energy balance and food intake^37,38^ as well as being linked previously with major depressive disorder^39^. *NEGR1* was associated with fluid intelligence score in our analysis which may raise concerns about the wider neurological safety of targeting this pathway.

Our results also highlight promising genes which may potentially make worthwhile targets. For example, predicted *SLC5A11* activation was highly associated with reduced risk of anthropometric (e.g. BMI) and cardiovascular traits but did not associate with any secondary potentially adverse effects in the PheWAS. SLC transporters are widely implicated in health and disease, and there is growing interest in strategies for therapeutic inhibition and activation of this protein family ^40^. Similarly, drugs targeting the *PRKCE* pathway may provide clinical benefit based on our analysis. *PRKCE* encodes PKC-ε, a member of the protein kinase C (PKC) serine/threonine protein kinases. The results of our PheWAS have predicted that *PRKCE* inhibition is not likely to elicit adverse secondary effects and may provide protection against high blood pressure. PKC-ε is known to be expressed in the heart and may confer a cardioprotective role during ischemic heart failure ^41^. Various *in vitro* and *in vivo* studies have provided evidence for a further role in mediating hypertrophy which may be dependent on *PRKCE* expression levels in the ischemic heart ^42^.

## Limitations

The study has several methodological limitations. Firstly, genetically predicted molecular traits represent the cumulative effect of lifelong exposure on the outcome and therefore cannot be used to directly predict the short-term benefit of a putative drug^43^. Secondly, the CVD outcomes derived from field 20002 in the UKB cohort are based on self-report. As such we firstly analyzed measured CVD risk factors from this cohort so that the risk of potential bias by implementing self-reported data from the population cohort is mitigated. Finally, our analysis is restricted to blood derived cis-eQTL due to availability of data, rendering them liable to loss of sensitivity to detect tissue-specific effects on disease susceptibility^44^. For example, the results of our analysis suggest that *PSRC1* expression at the 1p13.3 locus may associate with LDL cholesterol, although previous functional endeavors and those that using liver-derived expression data suggest that the likely causal gene for this signal is *SORT1* ^45,46^.

## Conclusions

The use of large-scale genetic studies employing Mendelian randomization can provide a cost-effective approach to accelerate the identification of viable drug targets to treat disease. Further pre-clinical studies are needed to validate the effectiveness of inhibiting targets which are prioritized by such efforts.

## Data Availability

Summary statistics used in this study are available from the eQTLGen consortium (https://www.eqtlgen.org/) and the MR-Base platform (http://www.mrbase.org/). All individual-level data analysed in this study can be accessed with an application to the UK Biobank study (https://www.ukbiobank.ac.uk/).

## Abbreviations

CVD: Cardiovascular disease
RCT: Randomized control trial
MR: Mendelian Randomization
GWAS: Genome wide association study
eQTL: expression quantitative loci
UKB: UK Biobank
PheWAS: Phenome wide association study
SNP: Single nucleotide polymorphism
BMI: Body mass index
DBP: Diastolic blood pressure
SBP: Systolic blood pressure
LDL: Low-density lipoproteins
TG: Triglycerides
IVW: Inverse variance weighted
GRS: Genetic risk score

## Acknowledgements

We are immensely grateful to the participants of the UK Biobank study whose data were accessed under application 15825 in this study. We would also like to thank Benjamin Elsworth for making the summary statistics publicly available from genome-wide association studies of cardiovascular risk factors for the benefit of this research. Finally, we wish to thank the efforts of the Neale Lab who conducted extensive heritability analyses in the UK Biobank which guided our selection of traits to analyze in our phenome-wide association analyses.

## Ethical approval

All individual participant data used in this study were obtained from the UK Biobank study who have obtained ethics approval from the Research Ethics Committee (REC - approval number: 11/NW/0382). All participants enrolled in UK Biobank have signed consent forms.

## Funding

This work was supported by the Integrative Epidemiology Unit which receives funding from the UK Medical Research Council and the University of Bristol (MC_UU_00011/4). GML is supported by the British Heart Foundation (FS/17/60/33474). TGR is a UKRI Innovation Research Fellow (MR/S003886/1).

### Disclosure

TRG receives research funding from GlaxoSmithKline and Biogen although neither company contributed to design or implementation of this study.

## Supplementary figures

**Supplementary Figure 1.**
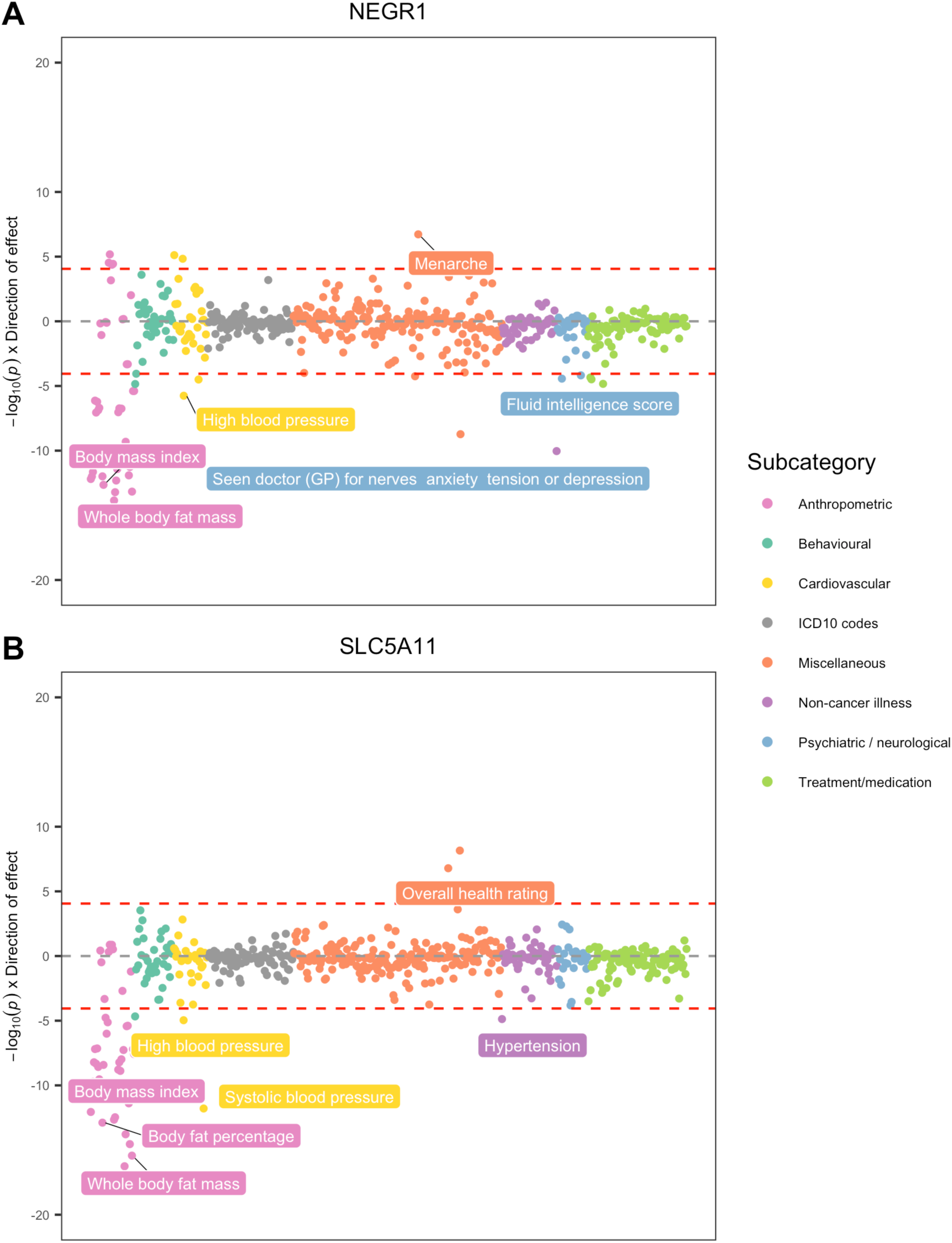
Phenome-wide association plot for predicted genetic inhibition of *NEGR1* (A) and *SLC5A11* (B) Each point on the plot represents the association between the respective genetic score and a complex trait in UKB. The y-axis indicates the -log^10^ p-value for the associations after orienting their direction of effect in line with predictive therapeutic treatment (i.e. statins have an LDL cholesterol lowering effect). Points are grouped and colored according to the corresponding subcategory for each trait. The horizontal dashed red line indicates the Bonferroni corrected threshold for multiple testing (-log10(0.05/(569)) = -log10(8.79×10^−5^).

